# Characterising the benefits and risks of vaccines approved through accelerated pathways

**DOI:** 10.1101/2025.05.31.25328649

**Authors:** Bethan Cracknell Daniels, Tim Endy, Joseph Lewnard, Jose Luis Di Fabio, Oscar Cortes-Azuero, Gabrielle Breugelmans, Simon Cauchemez, Alexander Precioso, Albert Ko, Rebecca E. Chandler, Stephen J. Thomas, Nicole Lurie, Henrik Salje

## Abstract

Alternative regulatory pathways for vaccine licensure without traditional Phase III efficacy trials are of increasing priority for epidemic pathogens. Using IXCHIQ, a chikungunya virus vaccine licensed through an accelerated pathway, we demonstrate that prior understanding of disease risk from infection can inform risk-benefit assessments for new vaccines. The mass deployment of IXCHIQ during an ongoing outbreak in La Réunion was suspended in those aged 65y+ following three deaths in vaccine recipients, although two appear unlikely to have been vaccine-linked. For individuals aged 18-64y, we find the vaccine benefits outweigh risks across epidemiological settings. In those aged 65y+, we estimate a net benefit in large outbreaks, which includes the La Réunion outbreak, and endemic settings. There is insufficient evidence of a net benefit in smaller outbreaks or in travellers. Our generalizable approach can help guide trial recruitment, inform vaccine implementation, and provide a foundation for weighing potential benefits against vaccine-associated risks.

## Background

Most vaccines are licensed based on the results of Phase III Randomised Placebo-Controlled Trials (RCTs), which assess vaccine safety and efficacy in reducing infection, morbidity and mortality. Planning such trials generally requires a predictable and detectable level of disease incidence. Alternative licensure pathways may therefore be needed for candidate vaccines against epidemic pathogens. For such pathogens, outbreaks can be sporadic, short-lived, and associated with broader public health emergencies—circumstances that each pose challenges for trial planning and implementation^1^. The US Food and Drugs Agency (FDA) and European Medicines Agency (EMA) therefore allow the use of immune correlates as a trial endpoint, where an immune marker is significantly associated with protection from a clinical endpoint^2^. These phase III safety and immunogenicity trials typically require fewer participants to determine immunogenicity than would be needed to detect clinical efficacy, as they instead measure the proportion of trial participants who mount a pre-specified immune response. While such trials are still designed to detect safety outcomes, including serious adverse events (SAEs), their smaller size limits the power to detect rare SAEs (<1/1000). Additionally, they may not capture safety signals in underrepresented subpopulations. As a result, the evidence generated may be insufficient for robust quantitative assessments of benefits and risks.

The recent licensure and rollout of the chikungunya vaccine IXCHIQ illustrates these challenges. IXCHIQ is a single-dose live attenuated vaccine developed by Valneva for preventing chikungunya disease^3^. Chikungunya virus (CHIKV) represents a major public health threat across global tropical and subtropical regions, causing disease manifestations ranging from acute febrile disease to encephalitis, long-term disabling arthralgia or arthritis, and death^4,5^. Age and comorbidities are risk factors for severe and fatal CHIKV infections^4–6^. While an estimated 34 million CHIKV infections and over 3,000 deaths occur each year, CHIKV outbreaks are unpredictable, explosive, and often short-lived, making Phase III RCTs difficult or even impossible to implement in most settings ^6,7^. The FDA and EMA licensed IXCHIQ in 2023/2024 based on a correlate of protection identified through passive transfer studies in non-human primates and confirmed in human seroepidemiological cohort studies ^8^. In the Phase III safety and immunogenicity pivotal trial (VLA1553-301), 263 of 266 vaccine recipients (98.9%) in the immunogenicity subset developed neutralising antibodies within 28 days, with responses sustained through three years of follow-up^3,9,8,10^. Among 3,082 IXCHIQ recipients followed for safety outcomes, 10 individuals (0.3%) developed chikungunya-like illness. Two vaccine-associated SAEs were reported in individuals aged 58 and 66 years. Although the trial reported no age-related differences in immunogenicity or safety, individuals aged 65+ represented only a small proportion of the trial population (n=463; 11%). The trial had a 94% probability of detecting at least one rare vaccine-associated SAE among the 3,000 vaccinated participants but was not designed to detect such events specifically in the higher-risk 65+ years age group.

The first mass vaccination with IXCHIQ began in April 2025 in response to an ongoing CHIKV outbreak in La Réunion, which began in August 2024. As of 21st May 2025, there have been 51,000 reported cases and 12 deaths associated with CHIKV infection, with 38 further deaths under investigation^11^. Mathematical modelling supporting the outbreak response estimates that 18–32% of the population have been infected (Simon Cauchemez, Personal communication). As part of epidemic control efforts, elderly individuals with comorbidities were initially prioritized for vaccination. An estimated 5,600 doses have been administered to those aged 65+ years (Valneva communication). However, three deaths have now been reported in vaccinated male individuals aged 77, 84, and 88, all of whom had multiple chronic comorbid conditions^12^. At the time of writing, these fatalities remain under investigation but only one death is likely associated with the IXCHIQ vaccine. However, limited representation of individuals aged 65+ years in the phase III trial meant there was insufficient evidence to confirm that the benefits of vaccination outweigh risks in this age group. As a result, the EMA has suspended IXCHIQ vaccination in individuals aged 65+ years, including in La Réunion^13^. The FDA also paused IXCHIQ use for individuals aged 60+ years until investigations of the SAEs reported from post-marketing use have been concluded.

The incidence of cardiovascular complications and blood clots from COVID-19 vaccines has motivated prior analytical exercises to quantify risk-benefit trade-offs^14,15,16^, but these efforts have been specifically focused on COVID-19 and have not assessed the strength of evidence for vaccine benefit. Here, we present a generalizable pathogen- and vaccine-agnostic framework for evaluating vaccine benefit-risk profiles that account for age-related variation in disease severity and vaccine-associated SAEs as well as uncertainty in SAE risk estimates when alternative licensure pathways are used ^17^. We then apply this model to IXCHIQ to help guide future vaccine implementation.

## Results

### Quantifying the risk and benefit of vaccines

We consider four key factors when comparing a vaccine’s risks and benefits: the risk of infection (defined as the attack rate, the proportion of an at-risk population that is infected), risk of a severe outcome (acute disease or death) following infection, vaccine efficacy against severe outcomes, and risk of vaccine-associated SAEs, including death. We define a vaccine-associated SAE as any untoward medical occurrence that results in death, is life-threatening, requires or prolongs hospitalization, or results in persistent or significant disability or incapacity following immunisation^18^. Specific population strata may experience differing adverse outcome risks from either infection or vaccination, for example, by age, sex, pregnancy status, and presence of chronic comorbid conditions, meaning that risk-benefit assessments could differ across subgroups. For example, if the risk of severe outcomes following infection is U-shaped by age (Figure S1A), as is common for many pathogens, but the risk of SAEs is age-independent, vaccine benefits will be greatest in infants and the elderly (Figure S1B). If, by contrast, the risk of SAEs also increases with age, greater benefits may shift to younger age groups (Figure S1C). The net benefit of vaccines also scales with attack rate and vaccine efficacy (Figure S1D-E).

We provide a generalized framework that firstly quantifies the risk of vaccine-associated SAEs that would be tolerated for different underlying risks of severe outcomes following infection, epidemiological settings, and vaccine efficacies (Figure 1). Secondly, the framework uses the number of SAEs observed to date to quantify the probability that the benefits of the vaccine outweigh the risks (Figure 2). We focus on four epidemiological scenarios in which a vaccine may be used, corresponding to a small outbreak (5% attack rate), a large outbreak (30% attack rate), an endemic setting (average annual infection of 2.4% over 10 years ^6^), and for travellers (10 days exposure risk during a large outbreak). For each scenario, we estimate the number of severe outcomes averted by a vaccine with 50% or 95% efficacy. The boundary between net benefit and net harm (dashed line) highlights the trade-off between risks of negative outcomes associated with infection and vaccination (Figure 1). As the risk of severe outcomes following infection increases (y-axis), net benefits persist (orange areas), even for vaccines associated with greater SAE risk (x-axis).

**Figure 1:**
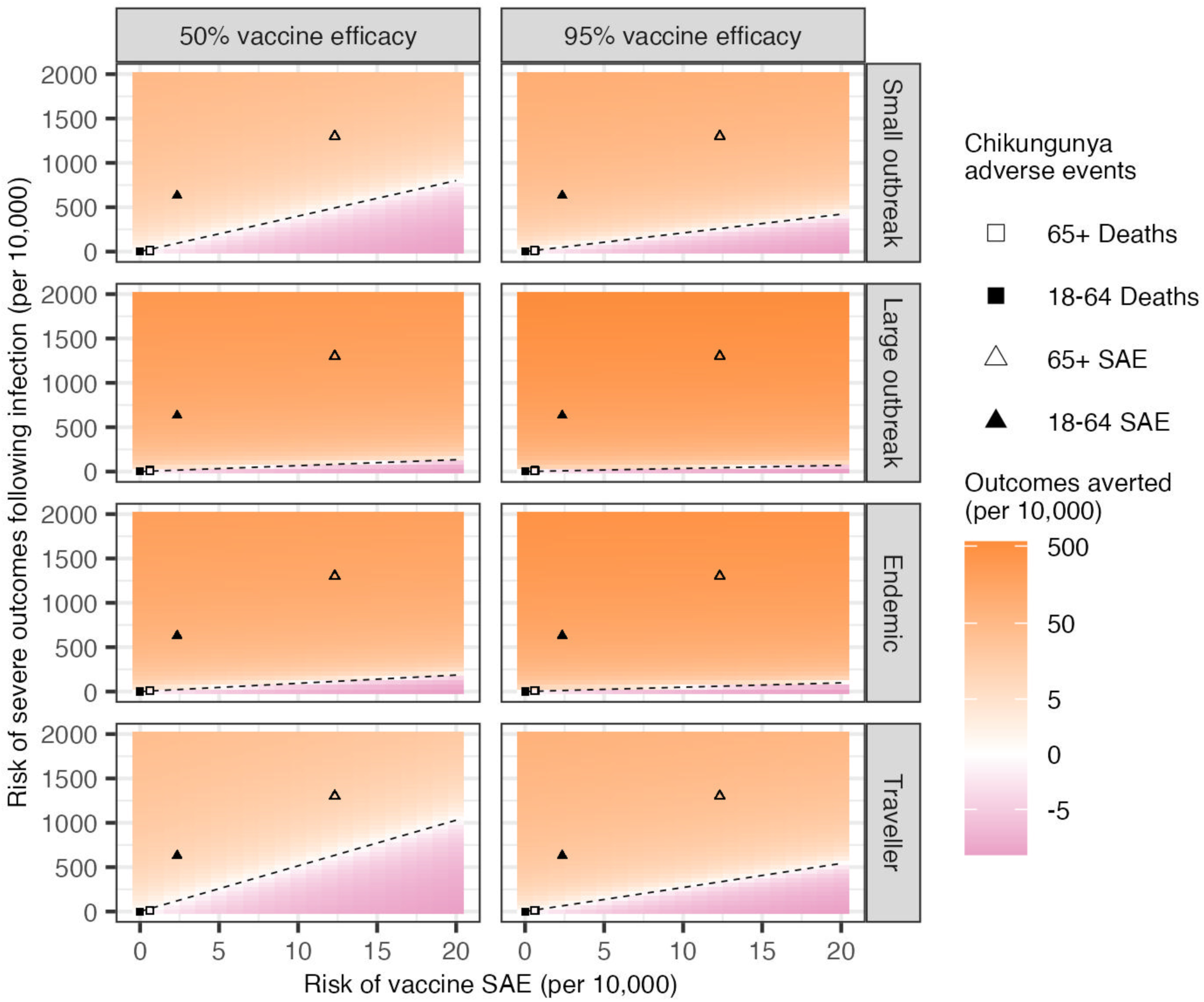
Impact of changes in the risk of vaccine SAEs or severe outcomes following infection on the number of cases averted across different epidemiological scenarios (rows), assuming 50% or 95% vaccine efficacy (columns). Positive and negative cases averted (per 10,000 in each age group) are coloured in orange and pink, respectively. The dashed diagonal line indicates the threshold of zero net benefit. The risks associated with chikungunya infection and vaccination for each age group (per 10,000) are denoted by the shapes. SAE: serious adverse event.

**Figure 2:**
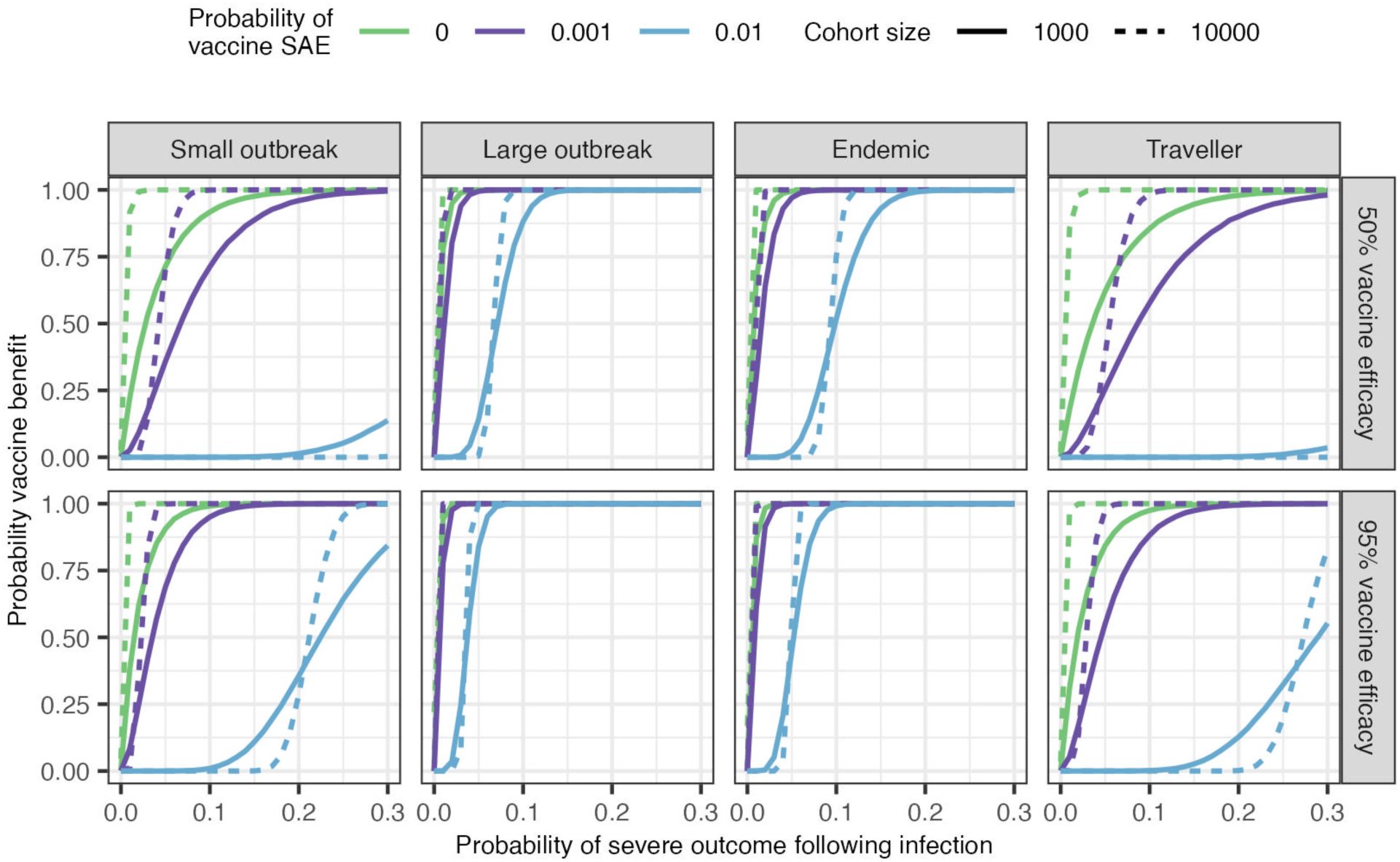
Evidence that benefits from vaccination outweigh risks. Probability that vaccine benefits outweigh the risks of vaccine SAEs assuming different probabilities of severe outcomes following infection, probabilities of adverse vaccine events and either 1,000 or 10,000 individuals vaccinated (cohort size). Estimates are shown for different epidemiological scenarios (columns), assuming 50% or 95% vaccine efficacy (rows).

We next assess the strength of the evidence that a vaccine’s benefits outweigh its risks, even in the absence of observed vaccine-associated SAEs. Specifically, we estimate the probability that a vaccine’s risk (number of SAEs observed in a given number of doses) is lower than its benefits (prevention of severe outcomes following infection), based on the number vaccinated to date, assumed efficacy, and the attack rate (Figure 2). To account for uncertainty in infection and vaccine risk estimates, we adopt a Bayesian approach that models both infection and vaccine risks as random variables. We show that, even if no SAEs are observed in 10,000 vaccinated individuals, the probability of vaccine benefit may still be <0.95 if the probability of severe outcomes following infection or the attack rate is low. Irrespective of vaccine-associated SAE risk, the epidemiological context (attack rate) strongly influences the vaccine benefit threshold. As the risk of severe outcomes following infection increases, vaccines remain beneficial despite higher vaccine risks, emphasizing that for severe diseases, vaccines are more likely to provide a net benefit. More generally, knowing vaccine efficacy and infection risk lets us establish upper bounds of acceptable vaccine risk. Conversely, for vaccines licensed without Phase III safety and efficacy RCTs, observed SAEs following real-world deployment can inform the minimum efficacy required to ensure benefit (Figure 2).

Given that vaccine-related deaths are a key concern for policymakers, but are often rare, we apply our framework to scenarios with no observed vaccine-associated deaths. Even without observed deaths, it may be unclear whether the benefits of preventing deaths outweigh the potential risks, especially in subpopulations with low vaccine coverage or when pathogen lethality is low. We show that for a highly lethal pathogen like Ebola virus (50% mortality following infection), even for a low attack rate (5%), having observed no deaths in 67 vaccinated individuals (assuming 95% efficacy) would allow us to conclude that vaccine benefits in preventing deaths outweigh the risks (Figure S2). For a less lethal pathogen, like SARS-CoV-2 in individuals with no immune protection (from prior infections or vaccination), observing no deaths in 3,400 vaccinated individuals would provide similar comfort.

### IXCHIQ vaccine as a case study

Considering the ongoing CHIKV outbreak in La Réunion and the suspension of vaccination for individuals aged 65+ years, we next applied our modelling framework to the IXCHIQ vaccine. During the phase III immunogenicity trial, two non-fatal vaccine-linked SAEs were reported—one in the 18–64-year age group (out of 2,736 vaccinated individuals) and one in the 65+ age group (out of 346 vaccinated individuals)^3^. As of April 30th, 2025, ∼37,500 IXCHIQ doses had been administered globally, including ∼16,000 doses in individuals aged 65+ years (Figure S3A) (Valneva communication). Among these vaccinated individuals, 25 non-fatal SAEs (5 and 20 in the 18-64 and 65+ year age groups, respectively), and one death linked to vaccination in a male individual aged 84 years have been reported (Figure S3B). A further two deaths post vaccination have been reported in two male individuals aged 77 and 88 years old, but the primary cause of death appears unlinked to the vaccine. Most non-fatal SAEs involved severe ‘chikungunya-like’ symptoms (consistent with the label), with at least 12/16 SAEs in La Réunion linked to IXCHIQ ^12^. We therefore include the 25 SAEs and 1 fatality in the main analysis. This gives as estimated 2.4 (95% confidence interval [CI]:0.8 - 5.5) non-fatal SAEs per 10,000 for ages 18–64 and 12.3 (95% CI:7.5 - 19.0) for those aged 65+, and 0.0 (95% CI:0.0 - 1.7) and 0.6 (95% CI: 0.02 - 3.4) death per 10,000, respectively (Figure S3C), indicating that the probability of SAEs and death following IXCHIQ vaccination are associated with age. Similarly, data from Paraguay and Brazil show that the risk of disease and death from CHIKV infection are strongly associated with age ^4–6^. Specifically, the infection fatality rate (IFR) is >20 times greater in those aged 65+ years than in 18-64 year olds, whilst the number of medically attended cases per 10,000 are 631.4 (95% CI: 627.3 - 635.5) and 1300.9 (95% CI: 1283.5 - 1318.3) in the same age groups (Figure S3C) ^4^.

Using our risk–benefit framework, we model the expected number of clinical chikungunya cases and deaths by age group and epidemiological scenario, both with and without vaccination. We assume 95% efficacy of IXCHIQ against severe outcomes and that vaccine SAEs are equivalent to medically attended chikungunya. We account for uncertainty in the risk of severe outcomes associated with both infection and vaccination (Figure 3). In all epidemiological scenarios, we find that vaccination results in fewer cases of medically attended chikungunya than SAEs, regardless of age (Figure 3A). The differences are greatest in large outbreaks in 65+ year olds (390 chikungunya cases without vaccination versus 20 cases and 13 SAEs with vaccination, per 10,000 individuals). Vaccination also leads to fewer deaths in large outbreaks and endemic settings, regardless of age group (Figure 3B). However, in small outbreaks or for travellers, vaccination may lead to a small excess risk of death in those aged 65+ (Figure 3D). Overall, an attack rate >5% confers net mortality benefits regardless of age group (Figure S4). As the attack rate in La Réunion is estimated at 18-32%, our analysis suggests that vaccination remains beneficial in reducing disease and mortality in all ages. This assumes that IXCHIQ efficacy is 95%, although this was not measured directly during the clinical trial. To ensure a net benefit against medically attended chikungunya, regardless of age or epidemiological context, efficacy must exceed 24% (Figure S5). Net mortality benefit in those aged 65+ requires efficacy above 16% in large outbreaks and 22% in endemic settings, whereas even a perfect vaccine does not provide net mortality benefit during small outbreaks or for travellers.

**Figure 3:**
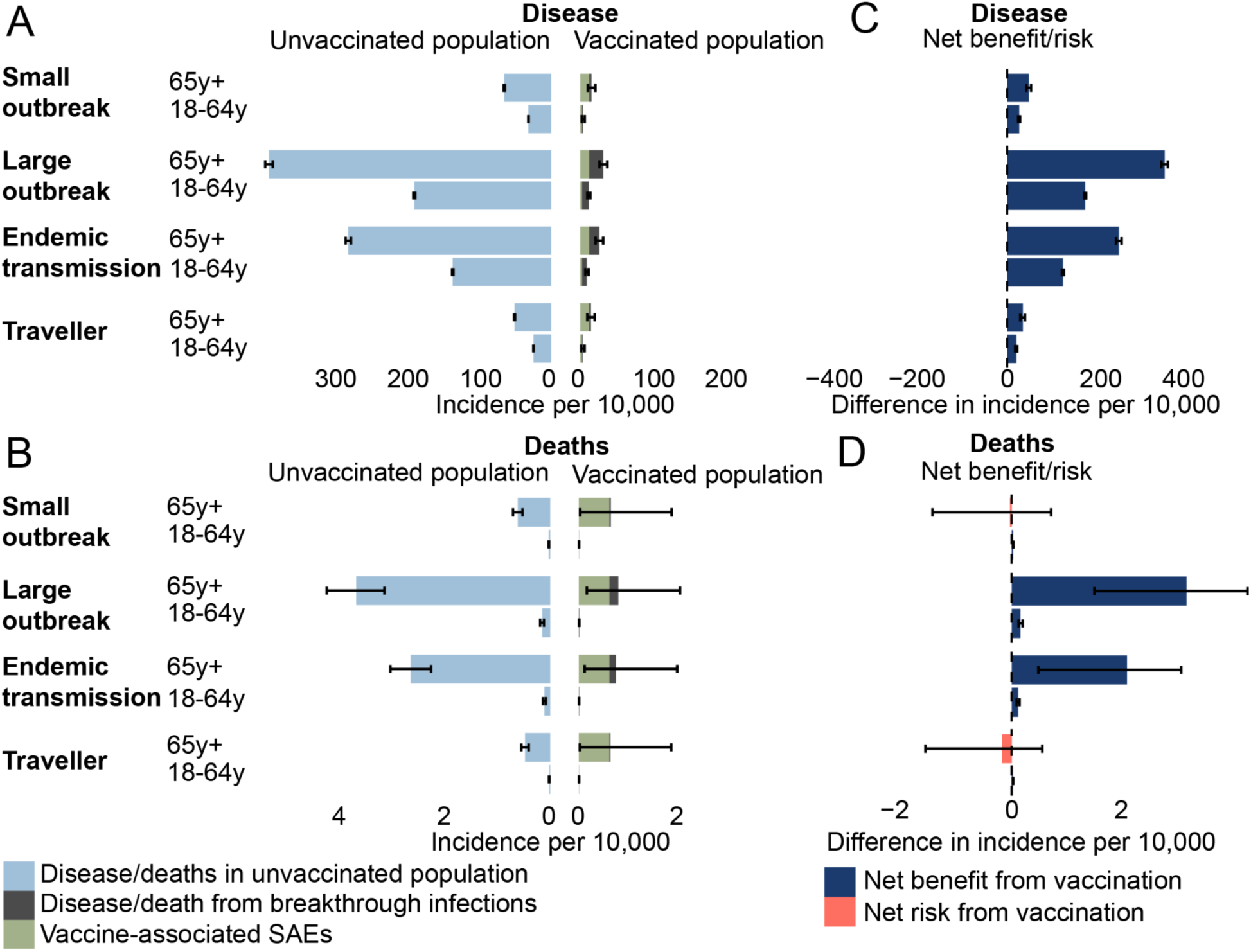
Risks and benefits of IXCHIQ vaccination assuming 95% vaccine efficacy. **(A)** Expected number of medically attended chikungunya cases / SAEs and **(B)** deaths per 10,000 individuals in each age group in the presence or absence of vaccination across different epidemiological scenarios and age groups. **(C)** Medically attended chikungunya cases / SAEs or **(D)** deaths averted across different epidemiological scenarios and age groups. Positive and negative cases averted (per 10,000 in each age group) are coloured in blue and red, respectively. See Methods for full details of assumptions. SAE: serious adverse event.

Next, we evaluate the strength of the evidence that IXCHIQ’s benefits outweigh its potential risks. Considering non-fatal SAEs first, we calculate the probability that the number of medically attended chikungunya cases averted by vaccination is greater than vaccine-associated SAEs, using both the data available at the end of the phase III immunogenicity trial and all the data available today (i.e., also including SAEs identified during mass vaccination in La Réunion) (Figure 4). We include uncertainty in both the number of SAEs observed and the underlying risk of severe acute disease from infection within a Bayesian framework. Assuming 95% vaccine efficacy, we find that at the end of the trial, there was a >0.95 probability that the vaccine’s benefit outweighed the risk of SAEs in all epidemiological scenarios in the 18–64 age group, and the large outbreak and endemic settings for the 65+ age group. Assuming 50% vaccine efficacy, we could still have concluded net benefit in both age groups in large outbreaks and endemic settings at trial completion (Figure S6). Using all the data available today, all age groups and epidemiological scenarios show vaccine benefit outweighing SAE risk, even if vaccine efficacy is 50% (Figures 4A, S6).

**Figure 4:**
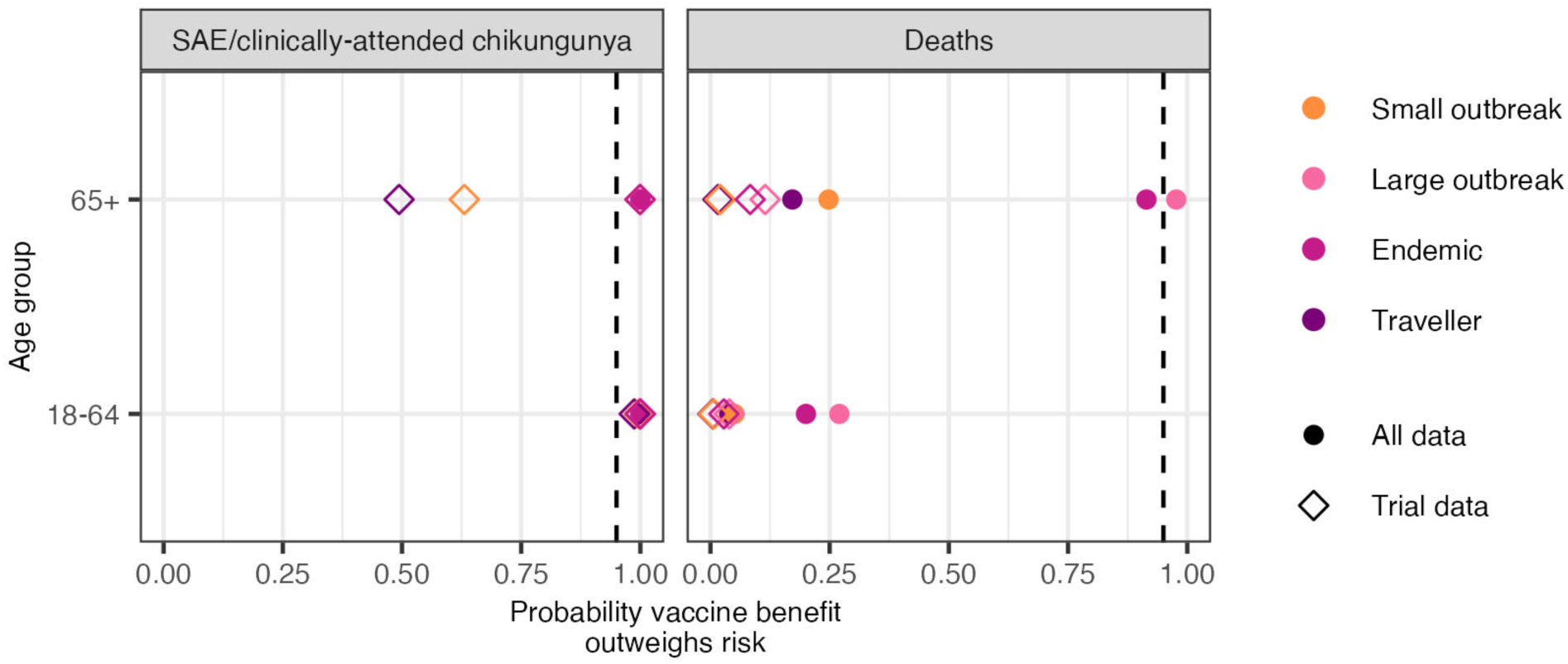
**Probability that the IXCHIQ vaccine benefits outweigh the risks of vaccine SAEs and vaccine deaths by age group and epidemiological scenario, assuming a vaccine efficacy of 95%**. Open squares show the evidence available at the end of the trial and solid circles show all evidence currently available. Vertical dashed line denotes 95% probability. SAE: serious adverse event.

Next, considering deaths, we find that at the end of the Phase III trial, there was insufficient evidence that the vaccine benefit outweighs the risk of death across all scenarios considered (Figure 4B). When we also include the data collected during mass vaccination, we find that despite the detection of one vaccine-associated death, the additional vaccine doses administered since the trial increase the probabilities of vaccine benefit against death for all scenarios, and there is >0.9 probability that the vaccine is beneficial in reducing deaths in the 65+ year age group in large outbreaks (0.98 probability of benefit) like La Réunion and endemic settings (0.91 probability of benefit). In sensitivity analyses, we find that if none of the observed deaths are linked to the vaccine, there is a >0.95 probability that the vaccine is beneficial in both large outbreaks and endemic settings. If all three observed deaths were linked, we still estimate a net benefit for those aged 65+ in large outbreaks and endemic settings, although the probability of vaccine benefit is <0.8 for all scenarios (Figure S7).

The lower evidence for net mortality benefits reflects the low IFR, especially for the 18–64 age group (Figure S3). We estimate that demonstrating a net mortality benefit in this age group during a large outbreak (30% attack rate) would require observing no deaths in >100,000 vaccinated individuals (Figure S2). This highlights that, for some pathogens or subpopulations, clinical trials may never generate sufficient evidence to conclusively show that benefits outweigh risks, despite the absolute risk remaining very low. For the 65+ year age group, no deaths in 4,570 vaccinated individuals would have demonstrated a net benefit.

Finally, there is not strict comparability between vaccine SAEs and medically attended chikungunya cases. Assuming instead that 50% of medically attended CHIKV infections are equivalent in health burden to SAEs (in line with chronic sequelae in half of medically attended cases ^19^), using all the data available today, we would still conclude a net vaccine benefit for all age groups and epidemiological scenarios (triangles moving down the y-axis in Figure 1, Figure S8). Alternatively, if fewer SAEs are ultimately linked to vaccination, the absolute net benefit would be greater than estimated here.

## Discussion

We have developed a generalizable, pathogen- and vaccine-agnostic framework to critically assess the risk–benefit trade-offs of vaccines licensed through alternative pathways, where risks are not fully characterized due to the absence of large Phase III safety and efficacy trials or the underrepresentation of high-risk subpopulations. This framework allows us to characterize both the absolute risk-benefits (cases averted) as well as the strength of the evidence (probability of benefit). We show that the underlying risk of infection and the subgroup risk of severe outcomes given infection are key to determining the settings where we can reliably conclude that vaccine benefits outweigh the risks. For IXCHIQ, vaccine benefits outweigh SAE risks across all ages and epidemiological scenarios when compared to medically attended or chronic chikungunya, consistent with the aim of CHIKV vaccines to reduce long-term morbidity, especially arthralgia.

Conversely, the evidence for IXCHIQ’s benefit against mortality depends on the age and epidemiological context. We estimate that the vaccine reduces the risk of death in the 18-64 age group in all epidemiological scenarios. However, the low IFR in this population means there is currently insufficient evidence to conclude that the mortality reduction outweighs the risks. It is only after observing no deaths in over 100,000 vaccinated individuals that it would be possible to conclude a benefit against mortality. In this context of very low absolute risk in the 18-64 age group, the clear net benefit of the vaccine in preventing severe disease should guide decision-making. In older individuals, where the absolute risk of mortality from both vaccine and infection is higher, our findings suggest that despite one probable death linked to vaccination, it remains beneficial in large outbreaks and endemic settings. Given the high attack rate estimated in La Réunion, this suggests that vaccination remained beneficial for mortality in those aged 65+. If all three deaths reported in vaccine recipients are ultimately found to be vaccine-related, we still estimate a net reduction in mortality risk from vaccination during large outbreaks, although the evidence for benefit is weaker. Critically, we did not account for comorbidities, which are strongly linked to the incidence of SAEs and were part of the vaccination campaign strategy developed by the authorities. Further, there have been more SAEs in males than females. With detailed disease and SAE data by comorbidity and sex, risk-benefit estimates could be refined to more specific subpopulations.

Our findings show that using existing understanding of the age specific risks from CHIKV infection, it would have been possible to conclude at the end of the Phase III trial (when ∼3,000 individuals had been vaccinated) that the benefits against clinical disease are greater than the risks from SAEs in the 18-64 age group in all epidemiological scenarios and older ages in scenarios with higher attack rates. Accounting for age and other factors associated with the risk of severe outcomes following infection during participant recruitment in vaccine trials can improve the ability to generate an evidence base of a vaccine’s risks and benefits. In the case of IXCHIQ, this would have involved recruiting more participants aged 65+ years, who comprised only 11% of trial participants despite being the population at greatest risk of severe disease following infection. Specifically, assuming a rare SAE (1 in 1000) and 95% efficacy, enrolling 4000 individuals aged 18–64 and 1000 aged 65+ years in the vaccine arm would have demonstrated net benefit across all epidemiological scenarios and age groups, when compared to medically attended chikungunya.

We show that the attack rate is a critical component in assessing a vaccine’s net benefit. In a new outbreak, this means decision-making must be supported by epidemic forecasting, anticipating the likely size of the epidemic ^20^. Typically, a public health response assumes a high attack rate to support aggressive vaccination, however, this may cause vaccine-associated harm in high-risk individuals if transmission is unexpectedly low. Conversely, a cautious approach to risk-benefit assessments, assuming a small outbreak, may underuse an effective vaccine if the outbreak escalates. Estimating pre-existing immunity through serosurveys and evaluating the likely impact of other control measures (e.g., vector control for CHIKV) is therefore essential. As CHIKV outbreaks become more frequent and vaccine-induced antibodies are broadly neutralising and expected to persist for years, vaccinated individuals are likely to remain protected in future outbreaks ^6,21^.

The lack of an RCT with clinical endpoints means that true vaccine efficacy estimates are unavailable through accelerated pathways. Our model can be used to identify the minimum vaccine efficacy necessary for the benefits of a vaccine to outweigh the risks. In the case of

IXCHIQ, we demonstrate that 25% vaccine efficacy would lead to a net benefit for medically attended chikungunya across all epidemiological scenarios and age groups. Conversely, even with 100% vaccine efficacy, individuals aged 65+ years in small outbreaks do not show a net mortality benefit. In general, however, we find that vaccine efficacy has less of an impact on determining the probability of vaccine benefit than the attack rate and risk of severe outcomes given infection.

The underlying probability of death from infection has only recently been quantified using a combination of death registry data and a seroprevalence study following a major outbreak in Paraguay^4^. Assuming an attack rate of 25% for the current outbreak in La Réunion, consistent with estimates from mathematical modelling supporting the outbreak response, we estimate approximately 225,000 infections to date in a population of 900,000. Based on the 12 confirmed deaths and 38 additional suspected deaths, this corresponds to an IFR ranging from 5 to 22 deaths per 100,000 infections, depending on how many of the suspected deaths are ultimately confirmed to be linked to CHIKV infection ^22^. This range is consistent with the ∼13 deaths per 100,000 found in Paraguay ^4^. Planned seroprevalence studies in La Réunion will help quantify the true infection burden in the territory, supporting a comparison of IFRs in different populations, which we have shown is vital for quantifying vaccine risks and benefits.

Our approach relies on necessary simplifications and assumptions. We did not account for indirect vaccine effects. An infection-blocking vaccine could confer herd immunity, benefiting unvaccinated individuals, including those ineligible, such as pregnant or immunocompromised people. Our estimates relied on IFRs from a single population and outbreak, and we assumed that medically attended CHIKV disease was approximately equivalent in disease burden to non-fatal vaccine-associated SAEs. Both vaccine-associated SAEs and medically attended CHIKV cases likely encompass substantial variability in symptom severity and healthcare-seeking behaviour. Nonetheless, varying the probability of severe disease from vaccination or infection consistently showed substantial vaccine benefit across scenarios and age groups. We also only considered two health outcomes (severe acute disease and death), ignoring some of the more severe infection outcomes. For example, encephalitis remains a poorly understood disease manifestation from CHIKV infection, especially in younger age groups, but has been consistently linked to infection ^23,24^. Protection from encephalitis, arthralgia and other chronic sequelae will be a further benefit from the vaccine not considered here, including economic impacts. Finally, we assume that a traveller is exposed to a high incidence outbreak for 10 days, however, an individual’s risk will depend strongly on their activities and use of protective measures.

Accelerated pathways are an important avenue for licensing vaccines where it is not feasible or practical to conduct Phase III safety and efficacy RCTs. In addition, it is faster, cost-effective and logistically/operationally preferable to do an immunogenicity study when possible, including for many non-epidemic pathogens, like pneumococcal vaccines ^25^. There must exist high confidence in vaccines approved through these mechanisms. We have shown that even when the safety of a vaccine is not fully characterized during licensure, we can quantify the strength of the evidence that the benefits of a candidate vaccine outweigh the risks and identify specific epidemiological scenarios and subpopulations that benefit from vaccination.

## Methods

Our analysis employs a risk-benefit framework to evaluate vaccine safety by comparing the expected harm prevented through vaccination against the potential harm caused by vaccine-associated SAEs. We first establish a simple model that calculates the number of severe outcomes expected in vaccinated versus unvaccinated populations under different epidemic scenarios, accounting for vaccine efficacy and baseline disease risk. From this, we derive a safety threshold—the maximum acceptable vaccine risk that still provides net benefit. We then implement this framework probabilistically using Bayesian inference to incorporate uncertainty in both disease and vaccine risk estimates, allowing us to calculate the probability that vaccination provides net benefit for different risk groups and outcomes.

### Quantifying the risk and benefit of vaccines

To understand the risks and benefits of a vaccine, we first estimate the risk of severe outcomes for vaccinated and unvaccinated individuals, given the underlying risk of infection, the risk of severe outcomes following infection, vaccine efficacy, and the risk of vaccine-associated SAEs. We assume that the risk of severe outcomes following infection and vaccine-associated SAEs varies by risk group (e.g., age). We characterise the underlying risk of infection through four scenarios: a small outbreak (5% attack rate), a large outbreak (30% attack rate), endemic transmission over 10 years (2.4% annual infection risk ^6^), and short-term exposure (10 days) of a traveller during a large outbreak.

The number of severe outcomes (𝑆) by vaccination status (𝑣 = vaccination, 𝑛𝑣 = no vaccination) by risk group 𝑎 is calculated as:

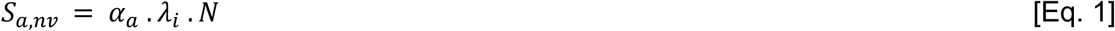

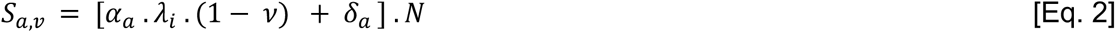

Where 𝑖 is the epidemiological scenario index, 𝜆_𝑖_ is the attack rate for scenario 𝑖, 𝛼_𝑎_ is the age-specific probability of severe outcome given infection, 𝛿_𝑎_ is the age-specific probability of vaccine–linked SAEs, 𝜈 is the vaccine efficacy against severe outcome, and 𝑁 is the population size.

The number of severe outcomes averted by vaccination is therefore:

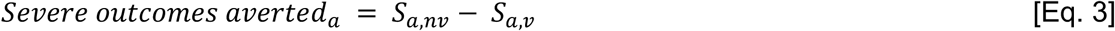

### Vaccine safety threshold

We evaluate a vaccine’s safety by comparing the observed risk of SAEs after vaccination to the expected background risk of severe outcomes given infection without vaccination. Using the same notation as above, to conclude vaccine benefit, we require that the total risk with vaccination does not exceed the risk without vaccination:

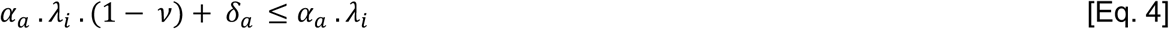

Rearranging, we obtain the maximum acceptable vaccine risk probability 𝛿_𝑚𝑎𝑥_ :

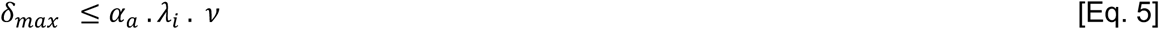

For vaccination to provide net benefit, we require 𝛿_𝑎_ ≤ 𝛿_𝑚𝑎𝑥_ .

### Probabilistic framework that benefits outweigh risks

To account for uncertainty in the infection and vaccine risk estimates, we adopt a Bayesian approach that treats both infection and vaccine risks as random variables. Specifically, we calculate the probability that the vaccine benefit threshold (𝛿_𝑚𝑎𝑥_ = 𝛼_𝑎_ . 𝜆_𝑖_ . 𝜈 ) exceeds the actual vaccine risk (𝛿_𝑎_).

For each adverse outcome (vaccine or infection) we use conjugate Bayesian inference. We assume a non-informative prior for the risks of severe outcomes given infection:

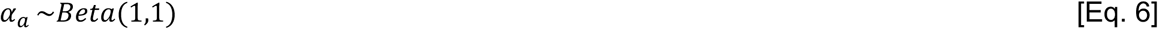

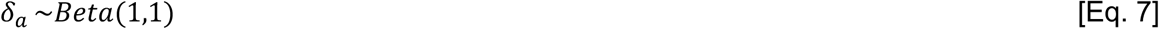

We assume that the number of adverse outcomes 𝑥 in 𝑛 total exposures (doses or infection) follow a binomial distribution:

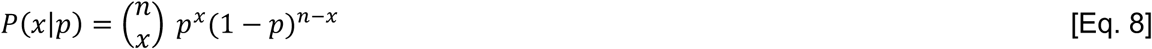

Where 𝑝 = 𝛼_𝑎_ when 𝑥 corresponds to adverse outcomes given infection, and 𝑝 = 𝛿_𝑎_ when 𝑥 corresponds to vaccine-linked SAEs.

The posterior distribution of adverse infection or vaccine-linked outcomes therefore follow a beta distribution:

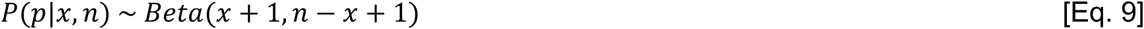

Drawing 10,000 samples from the posterior distributions of both 𝛼_𝑎_ (adverse infection outcomes) and 𝛿_𝑎_ (vaccine-linked SAEs), we calculate the probability of vaccine benefit as the proportion of samples where 𝛿_𝑎_ ≤ 𝛿_𝑚𝑎𝑥_. This represents the probability that vaccination provides a net benefit given the observed data.

### Power to detect at least one death from vaccination

Finally, we extended the safety threshold 𝛿_𝑚𝑎𝑥_ = 𝛼_𝑎_ . 𝜆_𝑖_ . 𝜈 to determine the minimum sample size required to conclude that the benefits of a vaccine in preventing death outweigh the risks, when no vaccine-associated deaths have been observed. Specifically, we calculate the sample size required to detect at least one vaccine-associated death with 80% power, assuming that the true vaccine risk is equal to the safety threshold 𝛿_𝑚𝑎𝑥_:

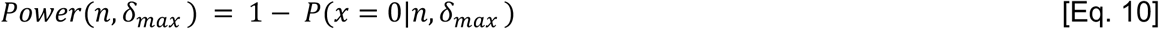

where 𝑥 ∼ 𝐵𝑖𝑛𝑜𝑚𝑖𝑎𝑙(𝑛, 𝛿_𝑚𝑎𝑥_ ).

### Application to the IXCHIQ vaccine

Next, we apply the framework described above to evaluate the IXCHIQ vaccine, analyzing two distinct severe outcomes: medically attended CHIKV cases and death. For each outcome, we estimate separate risk parameters for infection and vaccination by age group (18-64 and 65+ years).

### Data

*IXCHIQ* vaccine benefits are defined as the prevention of medically attended CHIKV cases (i.e. severe cases requiring hospitalisation or medical care) or deaths following infection. Estimates of the proportion of infections leading to medically attended cases and deaths were based on data from a large chikungunya outbreak in Paraguay (2022–2023), where a post-outbreak seroprevalence study was conducted (298 deaths, 142,412 medically attended cases, and 2.3 million infections) ^4^. The limited prior circulation of chikungunya allowed for a more accurate estimation of the infection fatality rate, as post-outbreak seroprevalence more closely reflected recent infections rather than background immunity. Medically attended cases were defined using national surveillance data and included symptomatic individuals who sought care and met the clinical case definition (sudden onset of fever and arthralgia or disabling arthritis not explained by another condition) ^4^.

*IXCHIQ* vaccine risk outcomes are SAEs (non-fatal or fatal). To compare vaccine risks and benefits, we assume that medically attended CHIKV cases are equivalent to a non-fatal SAE but vary this in a sensitivity analysis. We calculate the proportion of vaccine-associated SAEs stratified by age group (18-64, 65+ years) from data on the number of vaccine doses administered and the number of vaccine-associated SAEs ^12^ (Valneva communication).

### Statistical analysis

We estimate the risks and benefits of the IXCHIQ vaccine by calculating the net number of severe outcomes averted using Equation 3. To incorporate uncertainty in the observed data, we sample 10,000 times from binomial distributions representing the number of observed events (medically attended cases and deaths given infection, and fatal and non-fatal SAEs) given the total exposures (infections or vaccine doses). From these samples, we estimate the distribution of outcomes averted for each age group and epidemic scenario.

We also estimate the probability that vaccination provides net benefit for each outcome and age group separately. Using the posterior sampling approach (Equations 6-9), we draw 10,000 samples from the Beta distributions of infection risks (𝛼_𝑎_for deaths and cases) and vaccine risks (𝛿_𝑎_ for SAEs), then calculate the proportion of samples where 𝛿_𝑎_ ≤ 𝛿_𝑚𝑎𝑥_ for each combination of outcome (deaths or medically attended cases) and age group (18-64 or 65+ years).

We initially assumed a fixed vaccine efficacy of 95% for all age groups ^3^. We include sensitivity analyses where we assume vaccine efficacies ranging from 0-100%, attack rates of 0-50%, more or no vaccine-associated deaths, and that SAEs are instead comparable to chronic chikungunya cases ^19^.

## Data availability

All data is available at: https://github.com/bnc19/vaccine_risk_benefits.

## Code availability

Code is available at: https://github.com/bnc19/vaccine_risk_benefits.

## Data Availability

All data produced are available online at: https://github.com/bnc19/vaccine_risk_benefits

## Supplementary Figures

**Figure S1:**
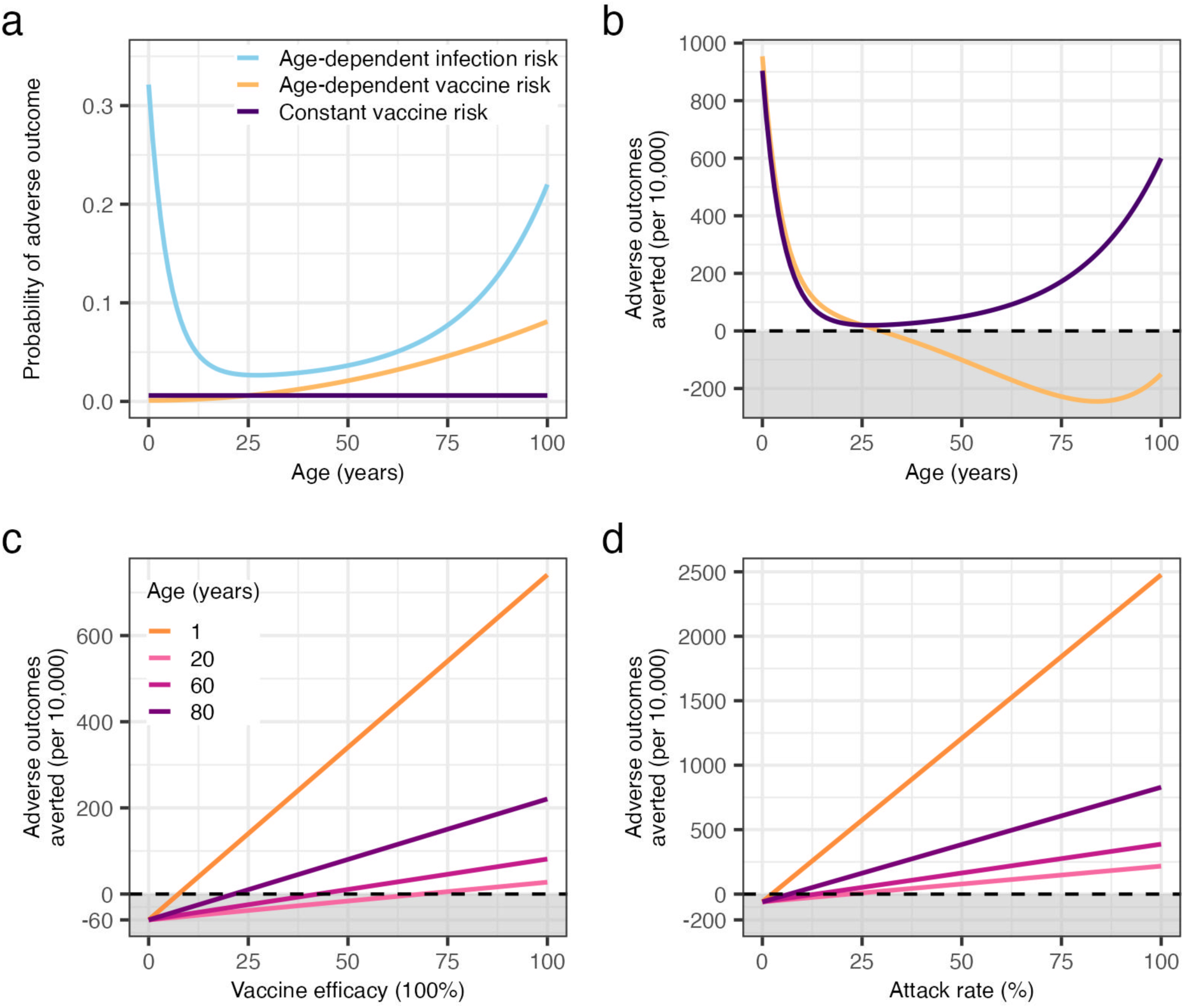
Factors affecting the risks and benefits of vaccination. **(A)** Age-specific probabilities of adverse age-dependent infection outcomes (blue), age-dependent adverse vaccine outcomes (orange), and age-independent adverse vaccine outcomes (purple). **(B)** Net adverse outcomes averted per 10,000 individuals by age under the two vaccine risk scenarios. **(C)** Adverse outcomes averted as a function of vaccine efficacy for individuals at different ages (1, 20, 60, 80 years), assuming a 30% attack rate. **(D)** Adverse outcomes averted as a function of the infection attack rate for individuals at the same four ages assuming 95% vaccine efficacy. The dashed horizontal line indicates the threshold of zero net benefit. Negative values (shaded region) indicate net harm.

**Figure S2:**
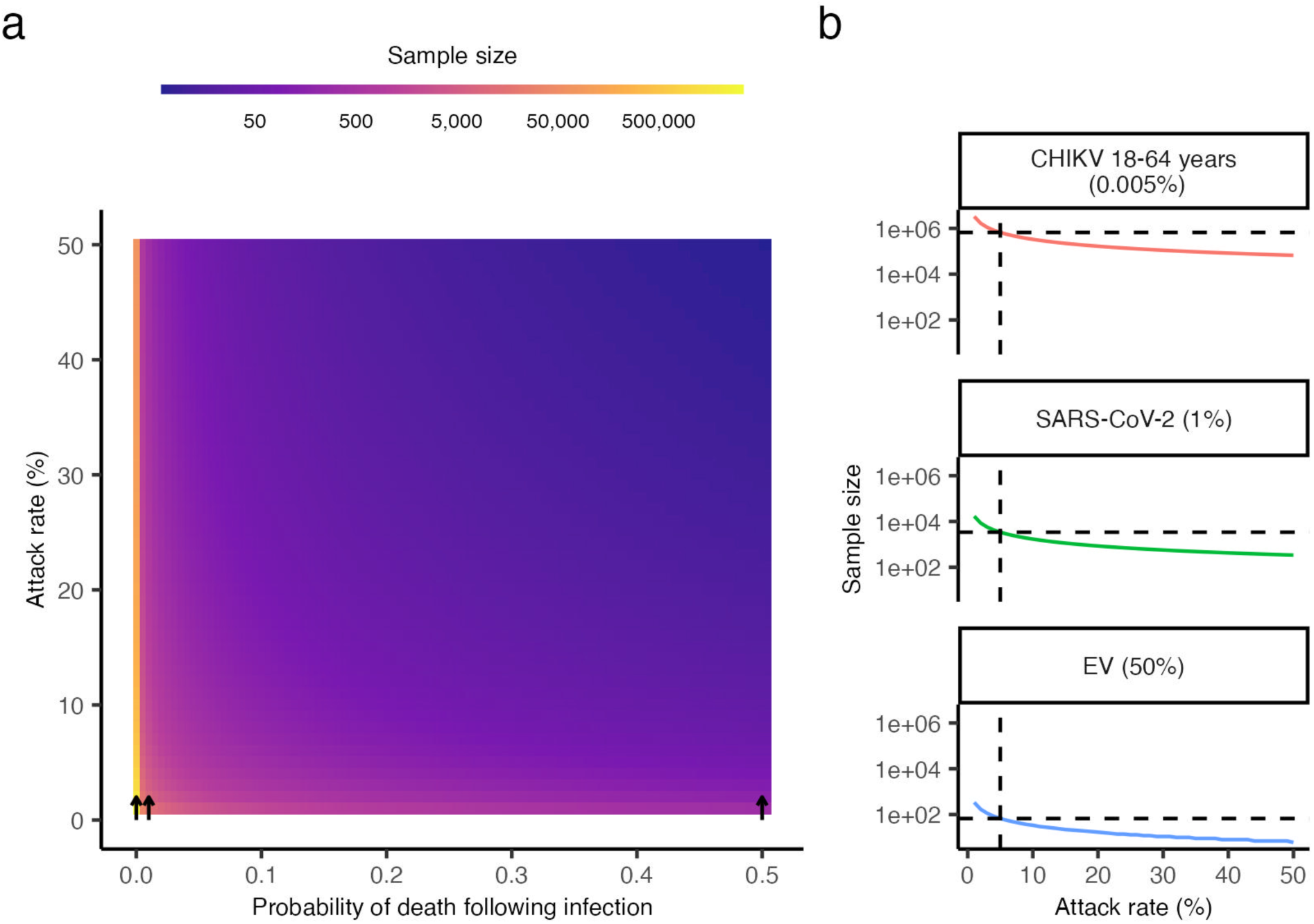
(A) Sample size needed to achieve 80% power to detect at least one death as a function of attack rate and probability of death from infection, assuming 95% vaccine efficacy. From left to right, the black arrows indicate the probability of death for CHIKV in the 18-64 age group (0.00005), SARS-CoV-2 (0.01), and EV (0.5). **(B)** Minimum population size required (vertical dashed line) for detecting that vaccine mortality risks do not exceed benefits with 80% power given a 5% attack rate (horizontal line), shown for the infection fatality rates (brackets) of CHIKV, SARS-CoV-2, and EV. CHIKV: Chikungunya virus. SARS-CoV-2: Severe acute respiratory syndrome coronavirus 2. EV: Ebola virus. SAE: serious adverse event.

**Figure S3:**
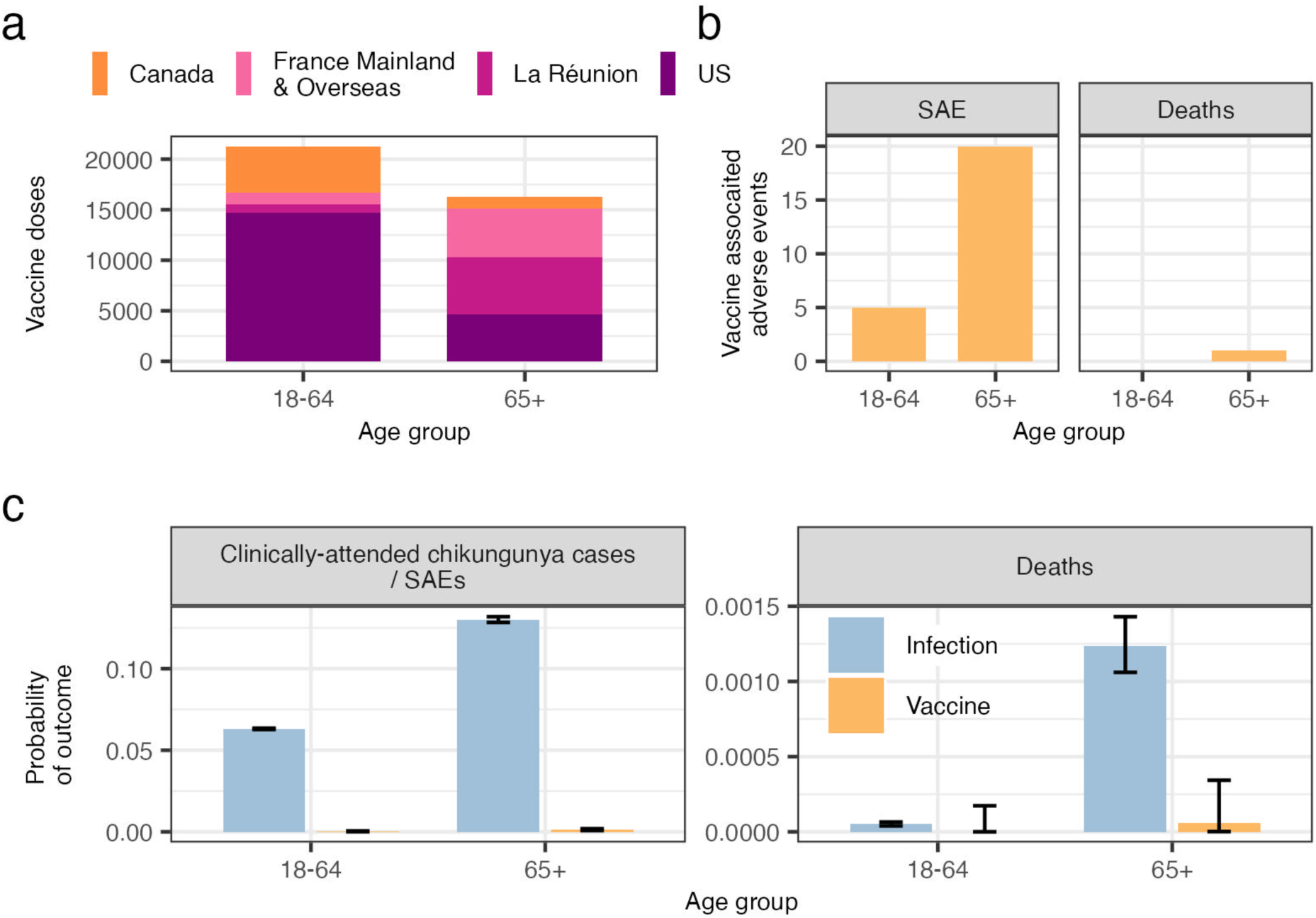
Risk of disease and death associated with chikungunya infection or IXCHIQ vaccination, stratified by age group. **(A)** Chikungunya vaccine doses administered as of April 2025 by country/territory. **(B)** Number of reported vaccine-associated SAEs and deaths. **(C)** Mean (bars) and 95% exact binomial confidence intervals (error bars) for the probability of medically attended chikungunya/SAEs and death associated with chikungunya infection (purple) or vaccination (yellow). SAE: serious adverse event.

**Figure S4:**
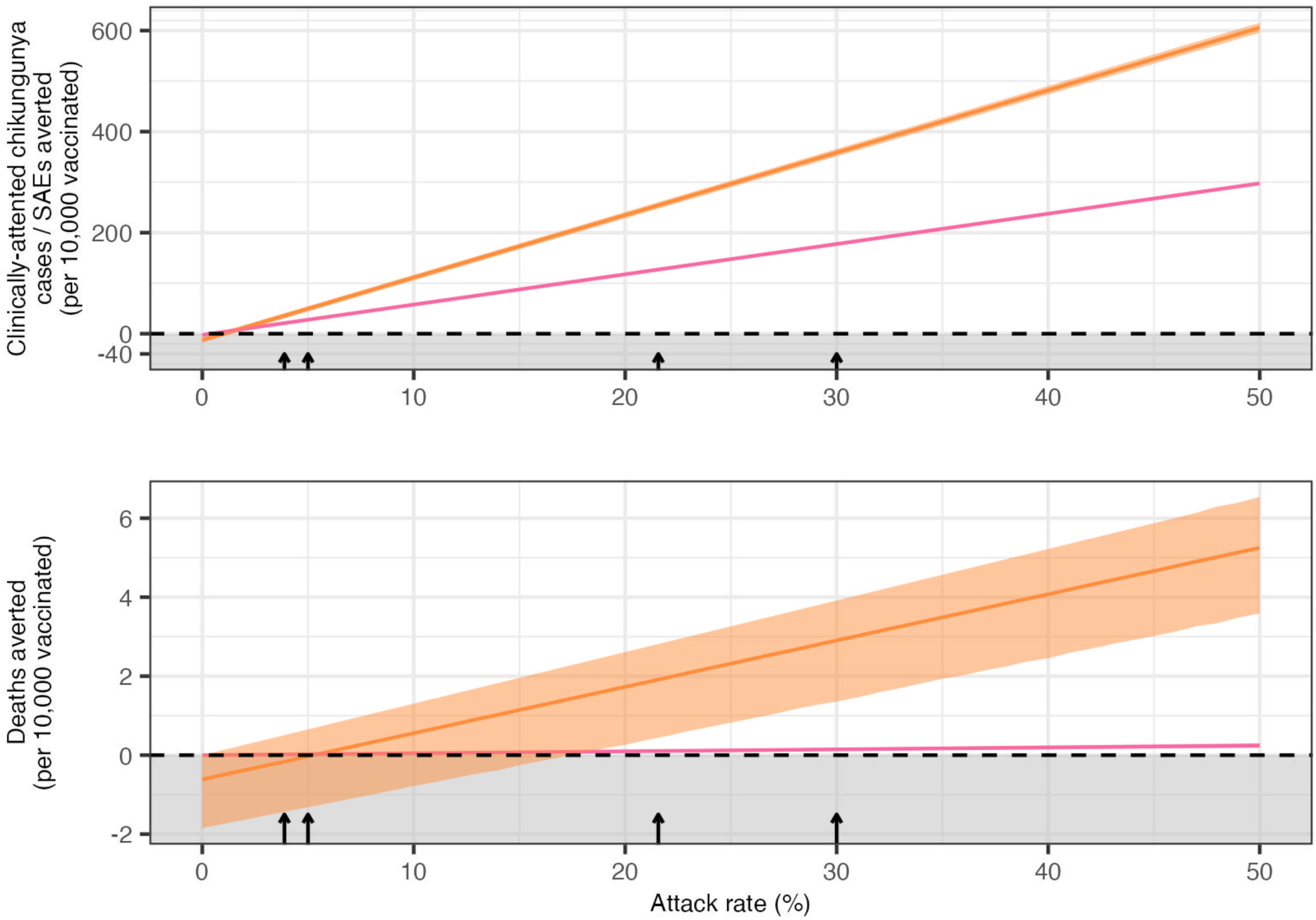
Impact of changes in the attack rate on cases averted by IXCHIQ vaccination. Number of medically attended chikungunya cases or deaths averted per 10,000 vaccinated in each age group (colors). The dashed horizontal line indicates the threshold of zero net benefit. Negative values (shaded region) indicate net harm. Arrows indicate epidemiological scenarios used in the main analysis, from left to right: traveller, small outbreak, endemic transmission, large outbreak. SAE: serious adverse event.

**Figure S5:**
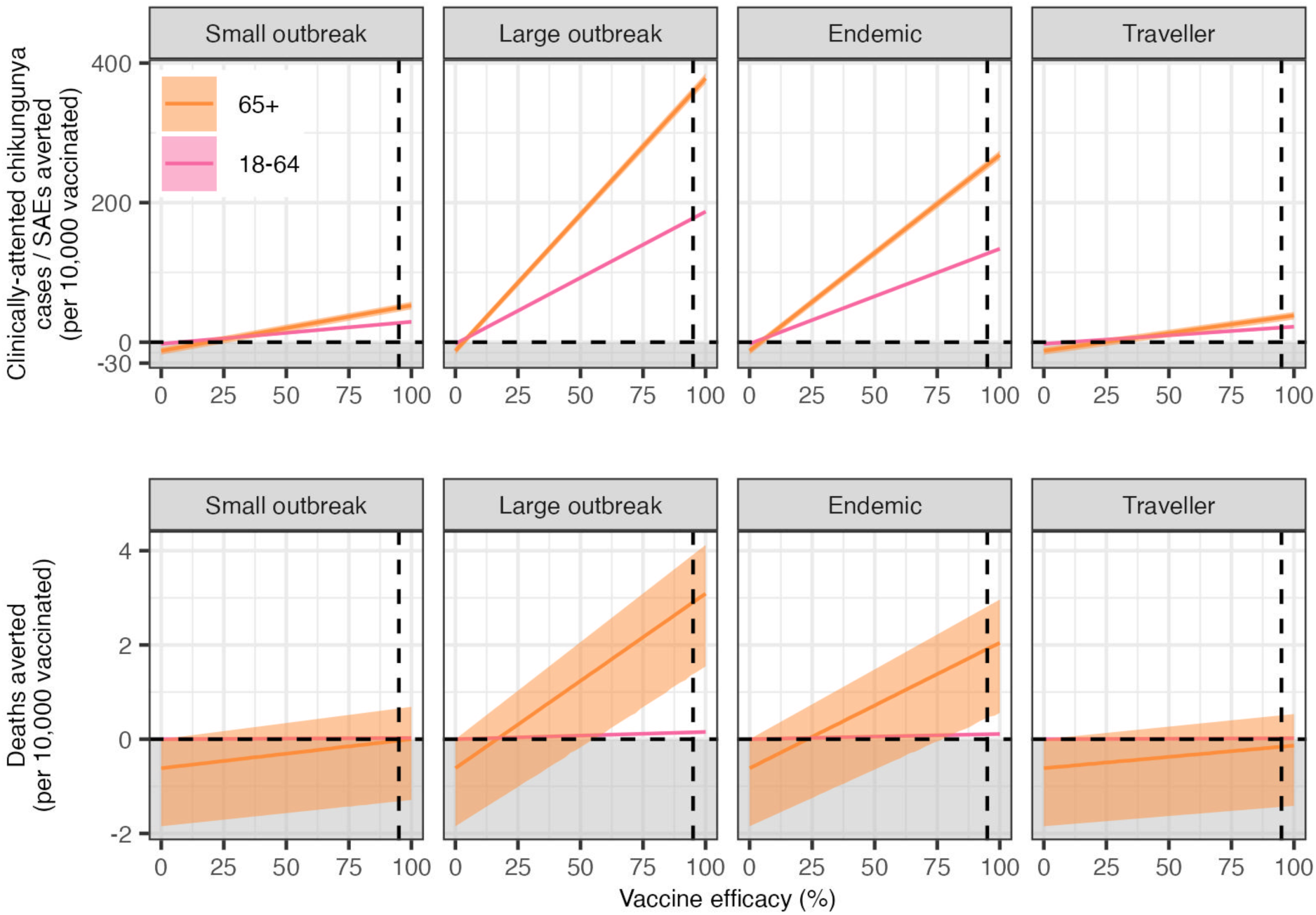
Impact of changes in vaccine efficacy on cases averted by IXCHIQ vaccination. Number of medically attended chikungunya cases / SAEs or deaths averted per 10,000 vaccinated (rows) in each age group (colors) by epidemiological scenario (columns). The dashed horizontal line indicates the threshold of zero net benefit. Negative values (shaded region) indicate net harm. Grey vertical line indicates baseline efficacy of 95%. SAE: serious adverse event.

**Figure S6:**
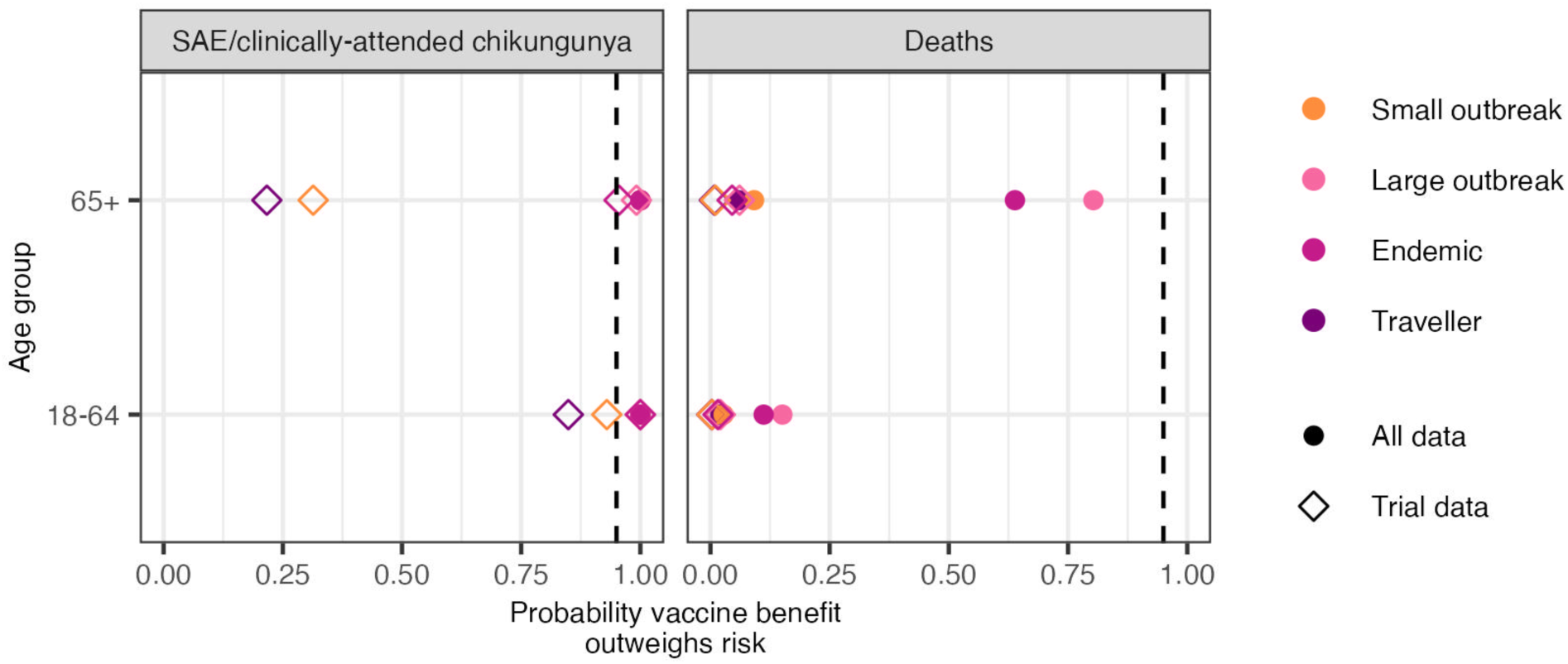
**Probability that the IXCHIQ vaccine benefits outweigh the risks of vaccine SAEs and vaccine deaths by age group and epidemiological scenario, assuming a vaccine efficacy of 50%**. Open squares show the evidence available at the end of the trial and solid circles show all evidence currently available. Vertical dashed line denotes 95% probability. SAE: serious adverse event.

**Figure S7:**
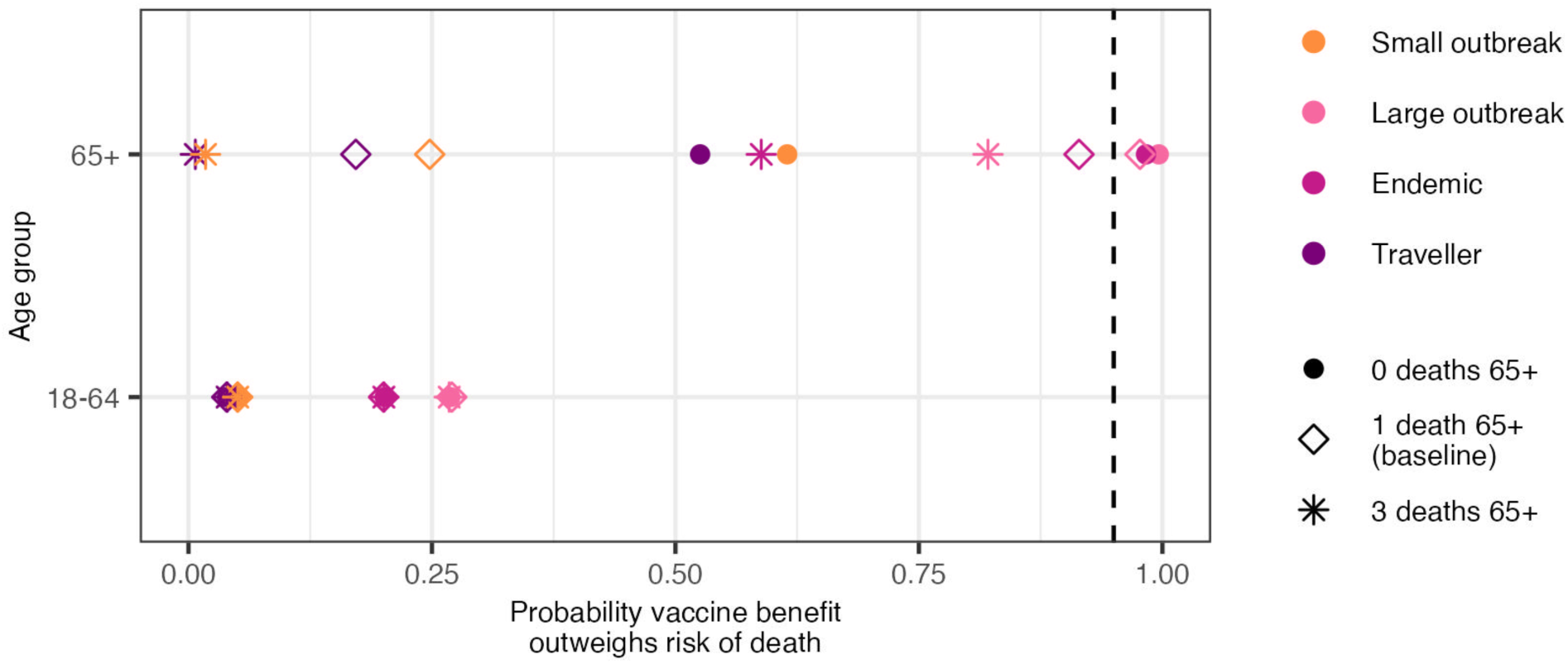
Probability that the IXCHIQ vaccine benefits outweigh the risks of vaccine deaths by age group and epidemiological scenario, using all the evidence currently available and assuming 0, 1 or 3 vaccine-linked deaths in the 65+ age group. Vertical dashed line denotes 95% probability.

**Figure S8:**
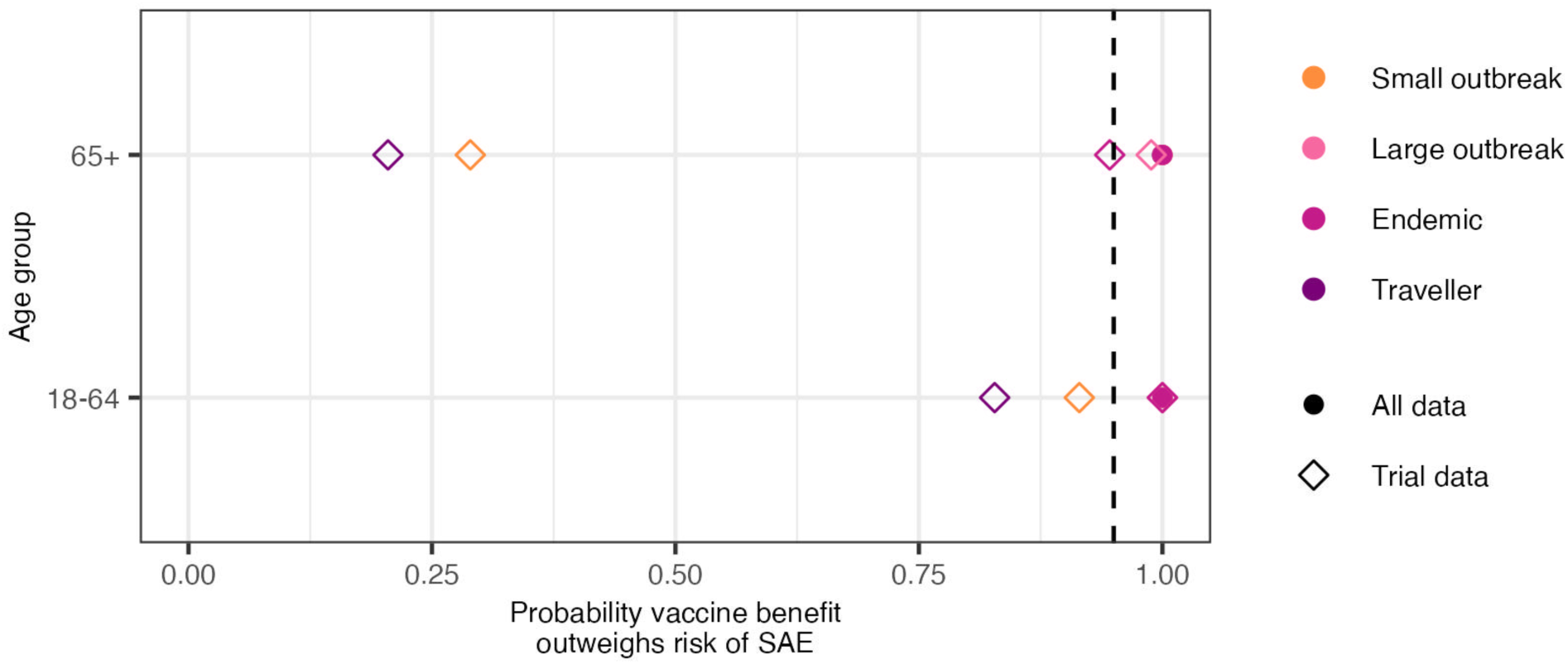
**Probability that the IXCHIQ vaccine benefits outweigh the risks of vaccine SAEs by age group and epidemiological scenario, assuming that 50% of medically attended CHIKV infections are equivalent in health burden to SAEs**. Open squares show the evidence available at the end of the trial and solid circles show all evidence currently available. Vertical dashed line denotes 95% probability. SAE: serious adverse event.

## References

1. Openshaw, P. J. M. Using correlates to accelerate vaccinology. Science 375, 22–23 (2022).

2. Liberti, L. et al. FDA facilitated regulatory pathways: Visualizing their characteristics, development, and authorization timelines. Front. Pharmacol. 8, 161 (2017).

3. Schneider, M. et al. Safety and immunogenicity of a single-shot live-attenuated chikungunya vaccine: a double-blind, multicentre, randomised, placebo-controlled, phase 3 trial. Lancet 401, 2138–2147 (2023).

4. Pérez-Estigarribia, P. E. et al. Modeling the impact of vaccine campaigns on the epidemic transmission dynamics of chikungunya virus outbreaks. Nat. Med. 1–7 (2025).

5. de Souza, W. M. et al. Spatiotemporal dynamics and recurrence of chikungunya virus in Brazil: an epidemiological study. Lancet Microbe 4, e319–e329 (2023).

6. Ribeiro dos Santos, G., et al. The global burden of chikungunya virus and the potential benefit of vaccines. medRxiv 2024.10.24.24315872 (2024) doi:10.1101/2024.10.24.24315872.

7. Rezza, G. & Weaver, S. C. Chikungunya as a paradigm for emerging viral diseases: Evaluating disease impact and hurdles to vaccine development. PLoS Negl. Trop. Dis. 13, e0006919 (2019).

8. Roques, P., et al. Effectiveness of CHIKV vaccine VLA1553 demonstrated by passive transfer of human sera. JCI Insight 7, (2022).

9. Valneva Reports Positive Three-Year Antibody Persistence Data for its Single-Shot Chikungunya Vaccine IXCHIQ®. Valneva https://valneva.com/press-release/valneva-reports-positive-three-year-antibody-persistence-data-for-its-single-shot-chikungunya-vaccine-ixchiq/ (2024).

10. Yoon, I.-K. et al. Pre-existing chikungunya virus neutralizing antibodies correlate with risk of symptomatic infection and subclinical seroconversion in a Philippine cohort. Int. J. Infect. Dis. 95, 167–173 (2020).

11. Santé publique France. Chikungunya in Reunion. Bulletin of May 21, 2025. https://www.santepubliquefrance.fr/regions/ocean-indien/documents/bulletin-regional/2025/chikungunya-a-la-reunion.-bulletin-du-21-mai-2025 (2025).

12. ANSM. Campagne vaccinale contre le chikungunya : point de situation sur la surveillance du vaccin Ixchiq (Chikungunya Vaccination Campaign: Update on Ixchiq Vaccine Monitoring). https://ansm.sante.fr/actualites/lansm-accompagne-la-campagne-vaccinale-contre-le-chikungunya-vaccin-ixchiq (2025).

13. EMA. EMA starts review of Ixchiq (live attenuated chikungunya vaccine). (2025).

14. Lau, C. L. et al. Risk-benefit analysis of the AstraZeneca COVID-19 vaccine in Australia using a Bayesian network modelling framework. Vaccine 39, 7429–7440 (2021).

15. Lewis, G. & Bonsall, M. Risk-benefit analysis of emergency vaccine use. Sci. Rep. 12, 7444 (2022).

16. Tran Kiem, C. et al. Benefits and risks associated with different uses of the COVID-19 vaccine Vaxzevria: a modelling study, France, May to September 2021. Euro Surveill. 26, (2021).

17. Patone, M. et al. Risk of myocarditis after sequential doses of COVID-19 vaccine and SARS-CoV-2 infection by age and sex. Circulation 146, 743–754 (2022).

18. World Health Organisation. Serious AEFI. https://www.who.int/groups/global-advisory-committee-on-vaccine-safety/topics/aefi/serious-aefi.

19. Kang, H. et al. Chikungunya seroprevalence, force of infection, and prevalence of chronic disability after infection in endemic and epidemic settings: a systematic review, meta-analysis, and modelling study. Lancet Infect. Dis. 24, 488–503 (2024).

20. Andronico, A. et al. Comparing the performance of three models incorporating weather data to forecast dengue epidemics in Reunion Island, 2018-2019. J. Infect. Dis. 229, 10–18 (2024).

21. Weber, W. C. et al. The approved live-attenuated Chikungunya virus vaccine (IXCHIQ®) elicits cross-neutralizing antibody breadth extending to multiple arthritogenic alphaviruses similar to the antibody breadth following natural infection. Vaccines (Basel) 12, 893 (2024).

22. Cerqueira-Silva, T. et al. Risk of death following chikungunya virus disease in the 100 Million Brazilian Cohort, 2015-18: a matched cohort study and self-controlled case series. Lancet Infect. Dis. 24, 504–513 (2024).

23. Sampaio, M. P. de S., et al. Detection of encephalitis-causing viruses reveals predominance of chikungunya virus in the state of Bahia, Brazil. Int. J. Infect. Dis. 145, 107090 (2024).

24. Bhardwaj, N., Kumar, V., Singh, P. & Kaur, J. Chikungunya encephalitis: A case report. J. Acute Dis. 13, 157–160 (2024).

25. King, D. F. et al. Realising the potential of correlates of protection for vaccine development, licensure and use: short summary. NPJ Vaccines 9, 82 (2024).

